# Bifurcation Left Main Stenting with or without intracoronary imaging: Outcomes From the EBC MAIN trial

**DOI:** 10.1101/2023.02.03.23285434

**Authors:** Annette Maznyczka, Sandeep Arunothayaraj, Mohaned Egred, Adrian Banning, Philippe Brunel, Miroslaw Ferenc, Thomas Hovasse, Adrian Wlodarczak, Manuel Pan, Thomas Schmitz, Marc Silvestri, Andrejs Erglis, Evgeny Kretov, Jens Flensted Lassen, Alaide Chieffo, Thierry Lefevre, Francesco Burzotta, James Cockburn, Olivier Darremont, Goran Stankovic, Marie-Claude Morice, Yves Louvard, David Hildick-Smith, the EBC MAIN (European Bifurcation Club Left Main Coronary Stent study) investigators

## Abstract

**Background:** The impact of intracoronary imaging on outcomes, after provisional versus dual-stenting for bifurcation left main (LM) lesions, is unknown.

**Objectives:** We investigated the effect of intracoronary imaging in the EBC MAIN trial (European Bifurcation Club Left Main Coronary Stent study).

**Methods:** 467 patients were randomised to dual-stenting or a stepwise provisional strategy. 455 patients were included. Intravascular ultrasound (IVUS) or optical coherence tomography (OCT) was undertaken at the operator’s discretion. The primary endpoint was death, myocardial infarction or target vessel revascularisation at 1-year.

**Results:** Intracoronary imaging was undertaken in 179 patients (39%; IVUS n=151, OCT n=28). As a result of IVUS findings, operators re-intervened in 42 procedures. The primary outcome did not differ with intracoronary imaging versus angiographic-guidance alone (17% vs. 16%; odds ratio (OR): 1.09 [95% confidence interval (CI): 0.66-1.82] p=0.738), nor for re-intervention based on IVUS versus none (14% vs.16%; OR: 0.86 [95% CI: 0.35-2.12] p=0.745). With angiographic-guidance only, primary outcome events were more frequent with dual versus provisional stenting (21% vs. 10%; OR: 2.24 [95% CI: 1.13-4.45] p=0.022). With intracoronary imaging, there were numerically fewer primary outcome events with dual versus provisional stenting (13% vs. 21%; OR: 0.54 [95% CI: 0.24-1.22] p=0.137).

**Conclusions:** In EBC MAIN, the primary outcome did not differ between patients who did or did not have intracoronary imaging. However, in patients without intracoronary imaging, outcomes were worse with a dual-stent than provisional strategy, and when intracoronary imaging was used, there was a trend toward better outcomes with the dual-stent than provisional strategy.

**Condensed abstract:** We investigated whether intracoronary imaging during LM bifurcation stenting was associated with less death, myocardial infarction and revascularisation at 1 year, for patients undergoing systematic dual versus stepwise provisional stenting. We included 455 patients from the EBC MAIN trial; 39% had intracoronary imaging. Overall, outcomes were similar between patients who did or did not have intracoronary imaging. In those with angiographic guidance only, outcomes were worse with dual versus provisional stenting (21% vs. 10%; OR: 2.24 [95% CI: 1.13-4.45] p=0.022). In those with intracoronary imaging, there was a trend toward better outcomes with dual versus provisional stenting (13% vs. 21%).

## Introduction

Left main (LM) bifurcation lesions may require demanding stenting techniques, with an increased risk of sub-optimal results(1). When angiography alone is used to guide percutaneous coronary intervention (PCI) it can be difficult to determine plaque calcification, atherosclerosis burden(2, 3) stent under-expansion and stent malapposition(4). Use of intravascular ultrasound (IVUS) or optical coherence tomography (OCT) may be associated with better stent expansion and fewer major adverse cardiac events (MACE) compared with angiography guidance alone (4-7).

Evidence from non-randomized studies(8-12) indicates that IVUS use in LM PCI is associated with lower mortality and that the benefit is most evident with complex lesions(8, 13). Moreover, the randomized ULTIMATE trial (Intravascular Ultrasound Guided Drug Eluting Stents Implantation in “All-Comers” Coronary Lesions), which included ∼55% LM lesions, showed lower rates of target vessel failure and stent thrombosis with use of IVUS. (14).

The EBC MAIN trial (European Bifurcation Club Left Main Coronary Stent Study) showed a trend to better clinical outcomes with provisional stenting compared to upfront dual-stenting for LM bifurcations(15). Our aim was to determine whether intracoronary imaging during LM bifurcation PCI was associated with less death, myocardial infarction (MI) and target vessel revascularization (TVR), for patients treated with a dual-stenting strategy versus provisional stepwise approach. We also aimed to determine the effect of intracoronary imaging on clinical outcomes, in relation to: extent of lesion calcification, lesion length in the side vessel, and stent length in the main vessel.

## Methods

### Trial Design

We performed a sub-analysis within the EBC MAIN trial. It was an investigator-led prospective, randomized, multicentre trial, involving 31 centres from 11 European countries. The trial was administered and overseen by the Cardiovascular European Research centre (CERC, Massy, France) and the study was supported by an unrestricted educational grant from Medtronic. The trial complied with the Declaration of Helsinki and the study protocol was approved by the relevant authorities in all countries involved in the study.

Details of the trial design and eligibility criteria have been published previously(15). In brief, from 2016 to 2019, patients who required PCI for unprotected bifurcation LM disease (Medina type 1,1,1 or 0,1,1) were randomised to a provisional single stent group, or to upfront dual-stenting. For patients in the systematic dual-stent group, the stent technique undertaken was at the operator’s discretion. In the EBC MAIN trial, 53% of patients randomized to the systematic dual-stent strategy had culotte stenting, 32% had T or TAP, 5% had DK Crush stenting, and 5% had no stent implanted in the side vessel. IVUS or OCT was undertaken at the operator’s discretion. The Onyx® zotarolimus-eluting stent (Medtronic) was used. Aspirin 75mg daily was continued long-term. Clopidogrel 75mg daily was given for at least 6 months (or appropriate dose of ticagrelor or prasugrel).

### Clinical outcomes

The clinical outcome data were adjudicated by the clinical events committee. The primary outcome was a composite of all-cause death, MI, and TVR at 1 year. The definitions for MI and TVR used in the trial have been described previously(15).

### Statistical analysis

Twelve patients were excluded from the analysis because of missing information on whether intracoronary imaging was undertaken (n=9), or because stenting of the left main was not performed (n=3). Normally distributed continuous data were summarized using mean ± standard deviation (SD). Categorical variables were reported as frequency and percentages. The trial endpoints were assessed using logistic regression. Regression models were used to assess treatment effects through interactions. We pre-specified the following sub-groups of interest according to procedural characteristics hypothesized to affect any association between intracoronary imaging use and the primary outcome: (i) bifurcation PCI strategy (dual-stent vs. provisional); (ii) lesion length in side vessel (<10mm vs. ≥10mm)(16); (iii) stent length in main vessel (<28mm vs. ≥28mm)(17) and (iv) angiographically defined extent of calcification (moderate-severe vs. none-mild)(16). All tests were assessed at the 5% significance level. There were no adjustments for multiple statistical comparisons. Statistics were performed using SPSS version 28.

## Results

### Population

The patient characteristics are shown in Tables 1 and 2. The flow of subjects through the study is shown in Figure 1. Of 455 patients included in this analysis, 226 patients were in the group randomized to the provisional stepwise approach, and 229 were in the group randomized to the systematic dual-stent approach. The mean age was 71.2 ± 9.8 years and 77% were male. The procedural indication was stable angina in 63%, and acute coronary syndrome in 37%.

**Table 1:** Population characteristics according to intracoronary imaging use, and by randomization to provisional or dual-stent strategy.

| Characteristic | Provisional Stepwise (n=226) |  |  | Systematic dual (n=229) |  |  | Overall |  |  |
| --- | --- | --- | --- | --- | --- | --- | --- | --- | --- |
|  | IVUS or OCT used (n=91) | Angiogram-guidance (n=135) | p-value | IVUS or OCT used (n=88) | Angiogram-guidance (n=141) | p-value | IVUS or OCT used (n=179) | Angiogram-guidance (n=276) | p-value |
| Age (years), mean ± SD | 70.8 ± 10.0 | 70.8 ± 10.3 | 0.981 | 69.2 ± 9.4 | 72.8 ± 9.4 | 0.027 | 70.4 ± 9.7 | 71.8 ± 9.9 | 0.132 |
| Male (%) | 76 (83.5%) | 102 (75.6%) | 0.151 | 63 (71.6%) | 107 (75.9%) | 0.470 | 139 (77.7%) | 209 (75.7%) | 0.636 |
| Ischemic symptoms (%) | 90 (98.9%) | 129 (95.6%) | 0.155 | 87 (98.9%) | 131 (92.9%) | 0.040 | 177 (98.9%) | 260 (94.2%) | 0.012 |
| Positive non-invasive imaging for ischemia (%) | 37 (40.7%) | 52 (38.5%) | 0.747 | 34 (38.6%) | 64 (45.4%) | 0.315 | 71 (39.7%) | 116 (42.0%) | 0.617 |
| Body mass index(kg/m2) mean ± SD | 28.6 ± 6.4 | 28.6 ± 4.8 | 0.956 | 28.9 ± 6.2 | 28.0 ± 4.8 | 0.199 | 28.8 ± 6.3 | 28.3 ± 4.8 | 0.341 |
| Diabetes (%) | 26 (28.6%) | 39 (28.9%) | 0.959 | 18 (20.5%) | 43 (30.5%) | 0.095 | 44 (24.6%) | 82 (29.7%) | 0.232 |
| Hypertension (%) | 72 (79.1%) | 107 (79.3%) | 0.980 | 73 (83.0%) | 114 (80.9%) | 0.689 | 145 (81%) | 221 (80.1%) | 0.806 |
| Hypercholesterolemia (%) | 67 (73.6%) | 91 (67.4%) | 0.317 | 67 (76.1%) | 96 (68.1%) | 0.191 | 134 (74.9%) | 187 (67.8%) | 0.104 |
| Current smoker (%) | 14 (15.4%) | 22 (16.4%) | 0.802 | 16 (18.2%) | 14 (9.9%) | 0.023 | 30 (16.8%) | 36 (13.0%) | 0.087 |
| Previous MI (%) | 26 (28.6%) | 34 (25.2%) | 0.572 | 31 (35.2%) | 35 (24.8%) | 0.091 | 57 (31.8%) | 69 (25.0%) | 0.111 |
| Previous PCI (%) | 38 (41.8%) | 55 (40.7%) | 0.879 | 37 (42.0%) | 60 (43.4%) | 0.856 | 75 (41.9%) | 116 (42.0%) | 0.978 |
| Previous stroke (%) | 8 (8.8%) | 8 (5.9%) | 0.410 | 8 (9.1%) | 8 (5.7%) | 0.324 | 16 (8.9%) | 16 (5.8%) | 0.200 |
| Peripheral vascular disease (%) | 10 (11.0%) | 21 (15.6%) | 0.328 | 10 (11.4%) | 26 (18.4%) | 0.152 | 20 (11.2%) | 47 (17.0%) | 0.085 |
| Renal failure (%) | 7 (7.7%) | 5 (3.7%) | 0.190 | 4 (4.5%) | 5 (3.5%) | 0.705 | 11 (6.1%) | 10 (3.6%) | 0.210 |
| Left ventricular function (%) |  |  | 0.148 |  |  | 0.562 |  |  | 0.582 |
| EF >50% | 59 (64.8%) | 84 (62.2%) |  | 58 (65.9%) | 82 (58.2%) |  | 117 (65.4%) | 166 (60.1%) |  |
| EF <50% | 28 (30.8%) | 25 (18.5%) |  | 23 (26.2%) | 39 (27.7%) |  | 51 (28.5%) | 64 (23.2%) |  |
| Unknown | 4 (4.4%) | 26 (19.3%) |  | 7 (8.0%) | 20 (14.2%) |  | 11 (6.1%) | 46 (16.7%) |  |
| Presentation (%) |  |  | 0.148 |  |  | 0.922 |  |  | 0.310 |
| Stable coronary disease | 54 (59.3%) | 94 (69.6%) |  | 53 (60.2%) | 84 (59.6%) |  | 107 (59.8%) | 178 (64.5%) |  |
| Acute coronary syndrome | 37 (40.7%) | 41 (30.4%) |  | 35 (39.8%) | 57 (40.4%) |  | 72 (40.2%) | 98 (35.5%) |  |
| Syntax score (%) |  |  | 0.008 |  |  | 0.765 |  |  | 0.036 |
| $\leq 22$ | 22 (24.2%) | 63 (46.7%) | | 29 (33.0%) | 57 (40.4%) | | 51 (28.5%) | 120 (43.5%) | |
| > 23 | 48 (52.7%) | 60 (44.4%) |  | 39 (44.3%) | 70 (49.6%) |  | 87 (48.6%) | 130 (47.1%) |  |
| Missing | 21 (23.1%) | 12 (8.9%) |  | 20 (22.7%) | 14 (9.9%) |  | 41 (22.9%) | 26 (9.4%) |  |
| Medina classification (%) |  |  | 0.310 |  |  | 0.484 |  |  | 0.225 |
| 1,1,1 | 84 (92.3%) | 119 (88.1%) |  | 80 (90.9%) | 124 (87.9%) |  | 164 (91.6%) | 243 (88.0%) |  |
| 0,1,1 | 7 (7.7%) | 16 (11.9%) |  | 8 (9.1%) | 17 (12.1%) |  | 15 (8.4%) | 33 (12.0%) |  |
| Adverse lesion features (%) |  |  |  |  |  |  |  |  |  |
| Trifurcation | 10 (11.0%) | 3 (2.2%) | 0.035 | 2 (2.3%) | 7 (5.0%) | 0.238 | 12 (6.7%) | 10 (3.6%) | 0.350 |
| Calcification ≥ moderate | 37 (40.7%) | 63 (46.7%) | 0.373 | 39 (44.3%) | 83 (58.9%) | 0.032 | 76 (42.5%) | 146 (52.9%) | 0.030 |
| Tortuosity ≥ moderate | 13 (14.3%) | 30 (22.2%) | 0.136 | 21 (23.9%) | 34 (24.1%) | 0.966 | 34 (19.0%) | 64 (23.2%) | 0.288 |
| Angle between LAD & LCx,<br>mean ± SD | 79.1 ± 18.1 | 81.2 ± 22.9 | 0.485 | 81.7 ± 17.7 | 82.9 ± 25.4 | 0.721 | 80.4 ± 17.9 | 82.0 ± 24.2 | 0.288 |

**Table 2:** Procedural characteristics according to intracoronary imaging use, and by randomization to provisional or dual-stent strategy.

| Characteristic | Provisional Stepwise (n=226) |  |  | Systematic dual (n=229) |  |  | Overall |  |  |
| --- | --- | --- | --- | --- | --- | --- | --- | --- | --- |
|  | IVUS or OCT used (n=91) | Angiogram-guidance (n=135) | p-value | IVUS or OCT used (n=88) | Angiogram-guidance (n=141) | p-value | IVUS or OCT used (n=179) | Angiogram-guidance (n=276) | p-value |
| Access site (%) |  |  | 0.939 |  |  | 0.982 |  |  | 0.993 |
| Femoral | 25 (27.5%) | 39 (28.9%) |  | 26 (29.5%) | 41 (29.1%) |  | 51 (28.5%) | 80 (29.0%) |  |
| Radial | 65 (71.4%) | 95 (70.4%) |  | 61 (69.3%) | 98 (69.5%) |  | 126 (70.4%) | 193 (69.9%) |  |
| Glycoprotein IIb/IIIa inhibitor (%) | 5 (5.5%) | 6 (4.4%) | 0.719 | 5 (5.7%) | 4 (2.8%) | 0.281 | 10 (5.6%) | 10 (3.6%) | 0.318 |
| Bivalirudin use (%) | 2 (2.2%) | 0 | 0.084 | 0 | 1 (0.7%) | 0.429 | 2 (1.1%) | 1 (0.4%) | 0.331 |
| Main vessel (%) |  |  | 0.532 |  |  | 0.379 |  |  | 0.110 |
| LM/ LAD | 72 (79.1%) | 102 (75.6%) |  | 70 (79.5%) | 105 (74.5%) |  | 142 (79.3%) | 207 (75.0%) |  |
| LM/ Cx | 19 (20.9%) | 33 (24.4%) |  | 18 (20.5%) | 36 (25.5%) |  | 37 (20.7%) | 69 (25.0%) |  |
| Stent length involving LM (mm), mean ± SD | 25.4 ± 14.3 | 25.6 ± 11.2 | 0.908 | 31.1 ± 18.5 | 32.3 ± 17.0 | 0.599 | 28.2 ± 16.6 | 29.0 ± 14.9 | 0.570 |
| Wire jailed after first stent (%) | 79 (86.8%) | 106 (78.5%) | 0.113 | 80 (90.9%) | 107 (75.9%) | 0.016 | 159 (88.8%) | 213 (77.2%) |  |
| TIMI flow in side vessel after 1<sup>st</sup> stent (%) |  |  | 0.268 |  |  | 0.014 |  |  | 0.015 |
| ≤ 2 | 3 (3.3%) | 9 (6.6%) |  | 0 | 8 (5.7%) |  | 3 (1.7%) | 17 (6.2%) |  |
| 3 | 88 (96.7%) | 126 (93.3%) |  | 80 (90.9%) | 102 (72.3%) |  | 168 (93.9%) | 228 (82.6%) |  |
| Missing data | 0 | 0 |  | 8 (9.1%) | 31 (22.0%) |  | 8 (4.5%) | 31 (11.2%) |  |
| Rewiring second vessel (%) |  |  | 0.443 |  |  | 0.578 |  |  | 0.393 |
| Yes | 84 (92.3%) | 128 (94.8%) |  | 83 (94.3%) | 136 (96.5%) |  | 167 (93.3%) | 264 (95.7%) |  |
| No | 7 (7.7%) | 7 (5.2%) |  | 2 (2.3%) | 1 (0.7%) |  | 9 (5.0%) | 8 (2.9%) |  |
| Missing | 0 | 0 |  | 3 (3.4%) | 4 (2.8%) |  | 3 (1.7%) | 4 (1.4%) |  |
| Stent to main vessel (%) | 91 (100%) | 135 (100%) |  | 88 (100%) | 141 (100%) |  | 179 (100%) | 276 (100%) |  |
| Stent to side vessel (%) | 18 (19.8%) | 33 (24.4%) | 0.633 | 85 (96.6%) | 133 (94.3%) | 0.436 | 103 (57.5%) | 166 (60.1%) | 0.797 |
| Reason for stenting side vessel in provisional group (%) |  |  | 0.528 |  |  |  |  |  |  |
| Dissection | 7 (7.7%) | 15 (11.1%) |  |  |  |  |  |  |  |
| Residual stenosis | 7 (7.7%) | 15 (11.1%) |  |  |  |  |  |  |  |
| Impaired flow | 1 (1.1%) | 0 |  |  |  |  |  |  |  |
| Other | 3 (3.3%) | 3 (2.2%) |  |  |  |  |  |  |  |
| Not applicable | 73 (80.2%) | 102 (75.6%) |  |  |  |  |  |  |  |
| Planned at start of the case to use IVUS (%) | 77 (84.6%) | 1 (0.7%) | <0.001 | 70 (79.5%) | 3 (2.1%) | <0.001 | 147 (82.1%) | 4 (1.4%) | <0.001 |
| OCT used (%) | 11 (12.1%) | 0 |  | 17 (19.3%) | 0 |  | 28 (15.6%) | 0 |  |
| IVUS used (%) | 80 (87.9%) | 0 |  | 71 (80.7%) | 0 |  | 151 (84.4%) | 0 |  |
| IVUS used in main vessel (%) | 45 (49.5%) | 0 |  | 19 (21.6%) | 0 |  | 64 (35.8%) | 0 |  |
| IVUS used in both vessels (%) | 35 (38.5%) | 0 |  | 52 (59.1%) | 0 |  | 87 (48.6%) | 0 |  |
| Operator reintervened with<br>balloon or stent as a result of<br>IVUS findings (%) | 28 (30.8%) | 0 |  | 14 (15.9%) | 0 |  | 42 (23.5%) | 0 |  |
| Length of lesion in LM (mm),<br>mean ± SD | 6.8 ± 3.3 | 6.02 ± 3.0 | 0.048 | 6.9 ± 3.0 | 6.02 ± 2.6 | 0.024 | 6.9 ± 3.2 | 6.0 ± 2.8 | 0.003 |
| Length of lesion in side branch<br>(mm), mean ± SD | 6.4 ± 4.8 | 5.4 ± 3.3 | 0.081 | 7.6 ± 5.2 | 8.1 ± 6.0 | 0.455 | 7.0 ± 5.0 | 6.8 ± 5.0 | 0.760 |
| Procedural duration (min),<br>mean ± SD | 86.8 ± 34.5 | 65.5 ± 33.4 | <0.001 | 94.3 ± 38.7 | 71.7 ± 37.1 | <0.001 | 90.5 ± 36.7 | 68.7 ± 35.4 | <0.001 |
| Fluoroscopy duration (min),<br>mean ± SD | 21.6 ± 11.9 | 20.5 ± 12.0 | 0.474 | 25.1 ± 18.4 | 22.8 ± 13.6 | 0.280 | 23.3 ± 15.5 | 21.7 ± 12.9 | 0.212 |
| X-ray dose (cGy.cm<sup>2</sup>) mean ± SD | 6682.0 ± 9189.7 | 7213.0 ± 5822.1 | 0.743 | 7404.7 ± 7091.5 | 7497.5 ± 6261.4 | 0.918 | 7138.8 ± 8206.4 | 7358.9 ± 6041.9 | 0.745 |
| Contrast volume (ml) mean ± SD | 218.1 ± 83.2 | 214.0 ± 97.1 | 0.743 | 239.2 ± 101.1 | 214.9 ± 92.1 | 0.062 | 228.5 ± 92.8 | 214.5 ± 94.4 | 0.120 |

**Figure 1.**
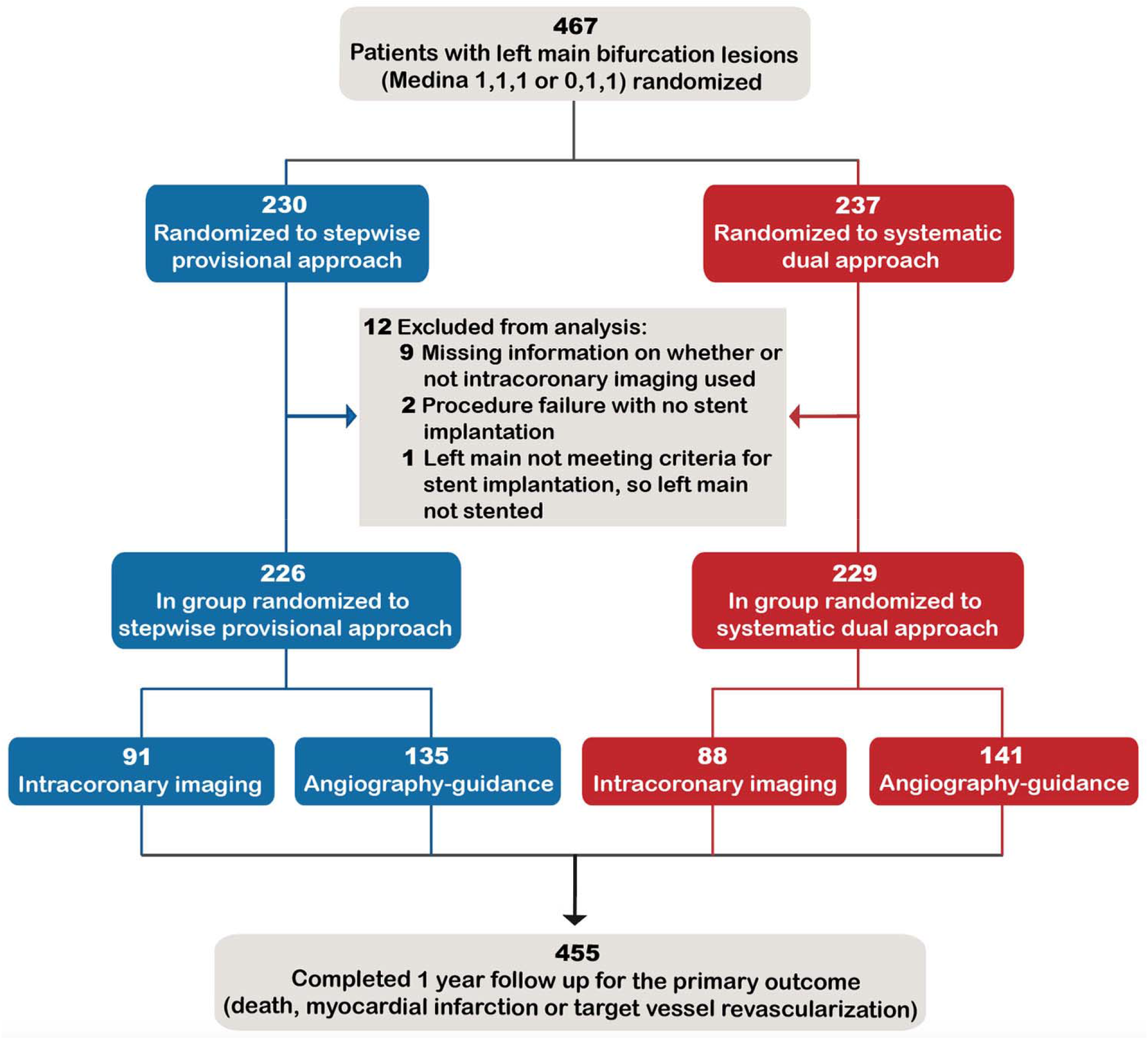
Study flow diagram. Number of patients included and excluded from the analysis.

Intracoronary imaging was used in 179 participants (39%; IVUS n=151, 33%; OCT n=28, 6%). The use of intracoronary imaging was similarly common with the stepwise provisional strategy (40%) as with the systematic dual-stent approach (38%). At the start of the case, operators planned to perform IVUS in 33% of procedures. Among the participants who had IVUS, 87 (58%) had both main vessel and side vessel imaged, and 64 (42%) had one vessel imaged. As a result of IVUS findings, operators reintervened with a balloon or stent in 42 procedures (28%), 28 of these procedures were in the stepwise provisional group, whereas 14 were in the systematic dual-stent group.

Lesions had severe coronary calcification in 98 participants (22%), and moderate calcification in 124 (27%). Calcified lesions (≥ moderately calcified) were less prevalent among patients who underwent intracoronary imaging than those who had angiography guidance only (42.5%, vs. 52.9%, p=0.030). In the group randomized to dual-stenting, there were again fewer calcified lesions in those who had intracoronary imaging than those who did not (44.3% vs. 58.9%, p=0.032), whereas in the provisional group there was no difference in lesion calcification between those who had intracoronary imaging or not (Table 2).

Notably, intracoronary imaging use was associated with longer procedure duration (90.5 ± 36.7 mins vs. 68.7 ± 35.4 mins, p<0.001), but was not associated with increased contrast volume, or radiation dose (Table 2). Furthermore, LM lesion length was on average longer in patients who underwent intracoronary imaging compared to those who had angiography guidance only (6.9 ± 3.2 vs. 6.0 ± 2.8, p=0.003).

### Clinical outcomes

The primary outcome (all-cause death, MI or TVR) occurred in 73 participants (16%) at 1 year. Overall, the primary outcome did not differ for patients who had intracoronary imaging versus angiography-guidance (17% vs. 16%; OR: 1.09 [95% CI: 0.66, 1.82] p=0.738).Similarly, the primary outcome did not differ for participants who had re-intervention based on IVUS findings compared to the rest of the participants (14% vs.16%; OR: 0.86 [95% CI: 0.35, 2.12] p=0.745).

There was a significant interaction between intracoronary imaging use and LM bifurcation PCI strategy, with respect to the primary outcome (p=0.009, Table 3). In those who did not have intracoronary imaging, the prevalence of the primary outcome was higher with the systematic dual approach compared to the stepwise provisional approach (21% vs. 10%; OR:2.24 [95% CI: 1.13, 4.45] p=0.022) (Figure 2). In those who had intracoronary imaging, there was a trend toward a lower prevalence of the primary outcome with the systematic dual approach compared to the stepwise provisional approach (13% vs. 21%; OR: 0.54 [95% CI: 0.24, 1.22] p=0.137). There was also a significant interaction between intracoronary imaging use and LM bifurcation PCI strategy with respect to MI and periprocedural MI (p=0.015 and p=0.035 respectively, Table 3). There were no interactions with intracoronary imaging use, the primary outcome, and the following: (i) extent of calcification; (ii) lesion length in the side vessel, or (iii) stent length in the main vessel (Table 4).

**Table 3:**
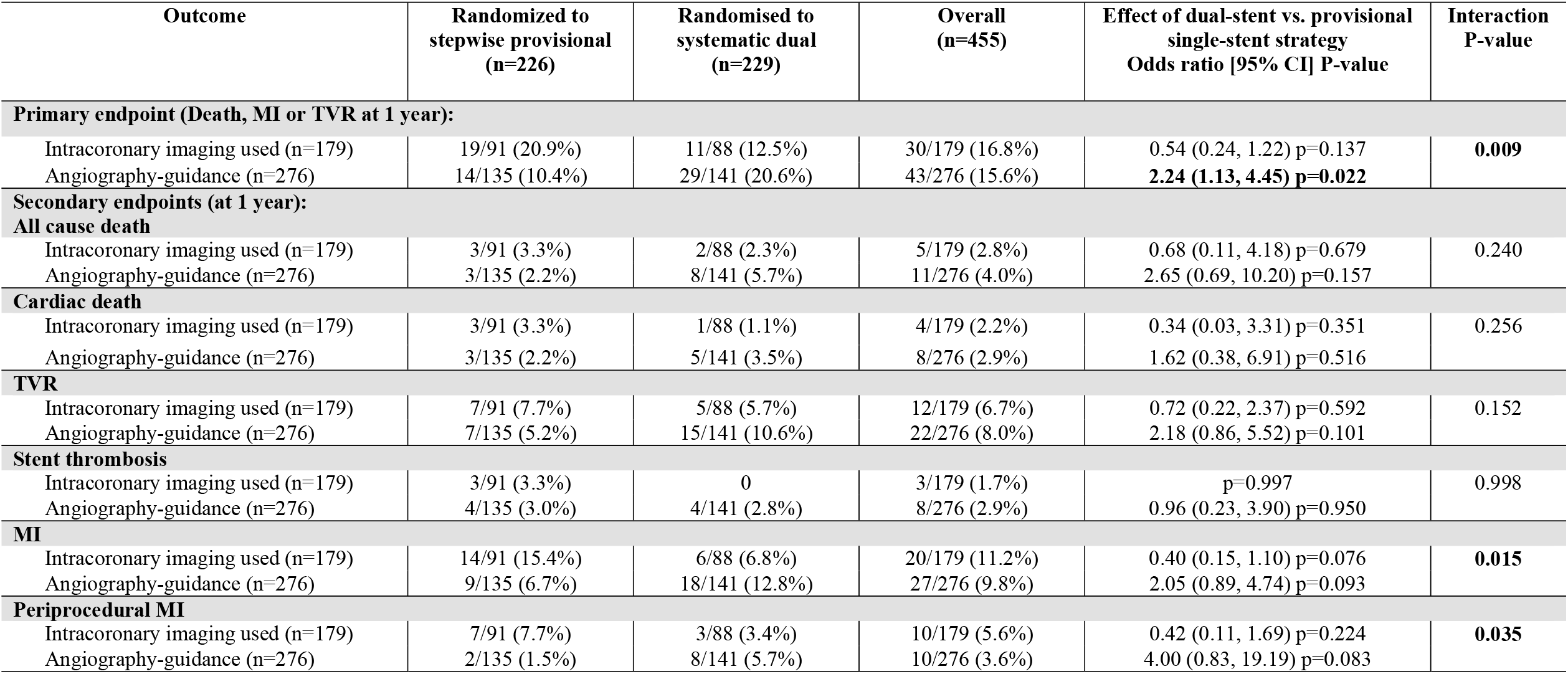
Clinical outcomes according to intracoronary imaging use, and by randomization to provisional or dual-stent strategy.

**Figure 2.**
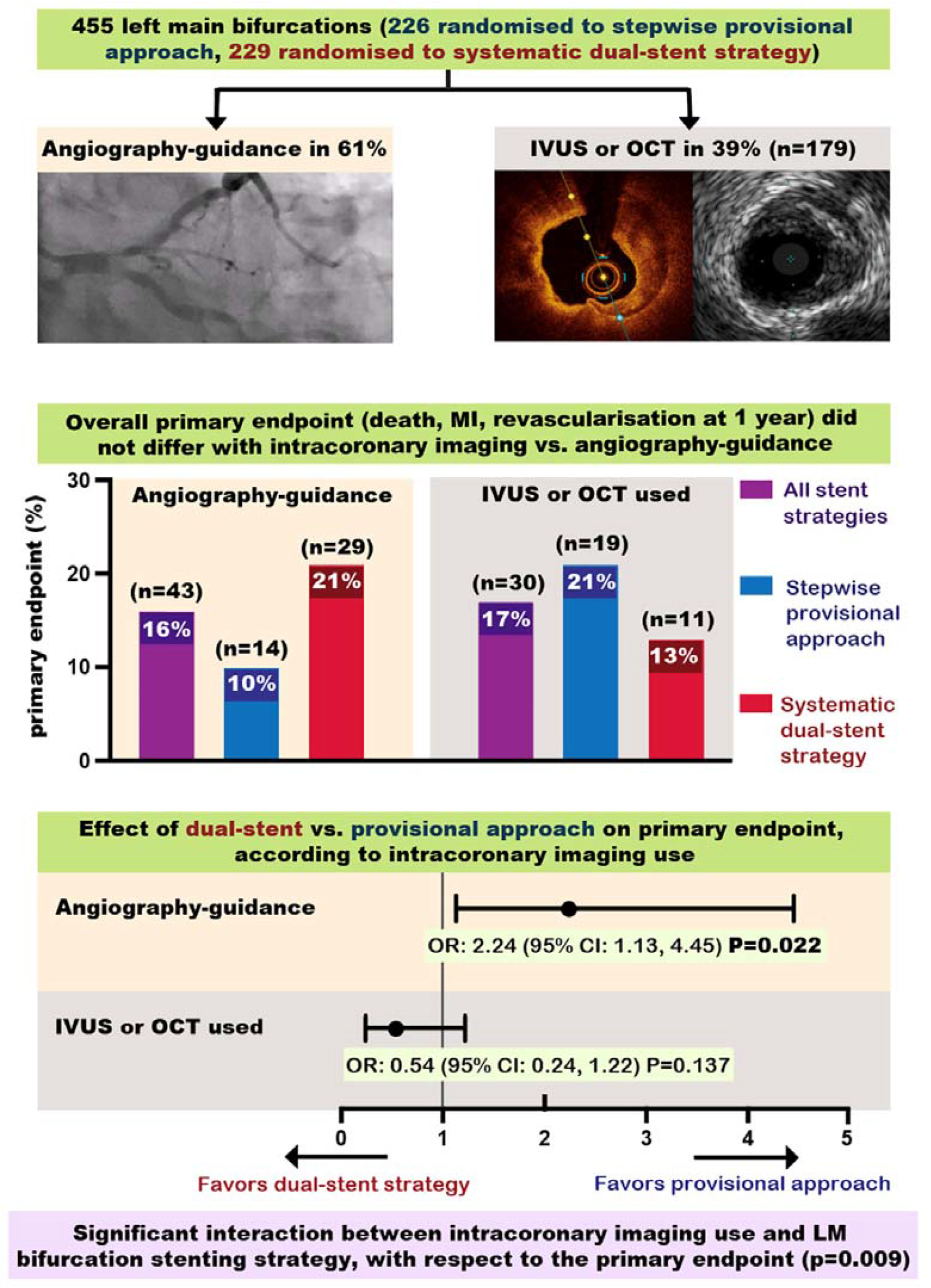
Summary of main findings (central illustration). Bar charts showing primary endpoint event rates and forest plots showing treatment effect estimates. Abbreviations: CI (confidence interval); IVUS (intravascular ultrasound); MI (myocardial infarction); OCT (optical coherence tomography); OR (odds ratio).

**Table 4:** Analysis of primary endpoint for intracoronary imaging use vs. angiography-guidance, according to potential confounding factors.

| Outcome | IVUS or OCT used<br>(n=event/total) | Angiography-guidance<br>(n=event/total) | Overall<br>(n=event/total) | Effect of intracoronary imaging use<br>vs. angiography-guidance<br>Odds ratio [95% CI] P-value | Interaction<br>P-value |
| --- | --- | --- | --- | --- | --- |
| (a) Lesion length in side vessel |  |  |  |  |  |
| < 10mm (n=374) | 26/147 (18%) | 32/227 (14%) | 58/374 (16%) | 1.31 (0.74, 2.30) p=0.350 | 0.960 |
| ≥ 10mm (n=76) | 4/28 (14%) | 11/48 (23%) | 15/76 (20%) | 0.56 (0.16, 1.97) p=0.366 |  |
| (b) Stent length in main vessel |  |  |  |  |  |
| < 28mm (n=258) | 18/106 (17.0%) | 27/152 (17.8%) | 45/258 (17%) | 0.95 (0.49, 1.83) p=0.871 | 0.525 |
| ≥ 28mm (n=197) | 12/73 (16.4%) | 16/124 (12.9%) | 28/197 (14%) | 1.33 (0.59, 2.99) p=0.493 |  |
| (c) Extent of calcification |  |  |  |  |  |
| Mild or none (n=233) | 17/103 (16.5%) | 19/130 (14.6%) | 36/233 (15%) | 1.16 (0.57, 2.35) p=0.692 | 0.854 |
| Moderate or severe (n=222) | 13/76 (17.1%) | 24/146 (16.4%) | 37/222 (17%) | 1.05 (0.50, 2.20) p=0.899 |  |

Missing data: length of obstruction in side branch =5 (IVUS or OCT used group =4, no IVUS or OCT group =1)

## Discussion

The main findings of our study are:

1. The primary outcome did not differ for patients who had intracoronary imaging vs. angiography guidance (17% vs. 16%; OR: 1.09 [95% CI: 0.66, 1.82] p=0.738).
2. The primary outcome did not differ for participants who had re-intervention based on IVUS findings compared to the rest of the participants (14% vs.16%; OR: 0.86 [95% CI: 0.35, 2.12] p=0.745).
3. In patients where no intracoronary imaging was undertaken, primary outcome events were more frequent with the systematic dual-stent strategy compared to the provisional approach.
4. When intracoronary imaging was used, there was a trend toward fewer primary outcome events with the systematic dual-stent strategy compared to the provisional approach.

These findings are not uniform, but suggest that intravascular imaging may be important to optimize the results from a dual-stent strategy during LM bifurcation PCI. This is in accordance with previous non-left-main bifurcation studies which have shown that IVUS guidance for bifurcations treated with a dual-stent technique was associated with lower rates of MI and late stent thrombosis than angiographic guidance(18), and that IVUS guidance for bifurcation PCI was associated with lower rates of cardiac death and MI, compared to angiography-guidance(19).

Among participants who underwent the stepwise provisional approach, we unexpectedly observed more primary outcome events when intracoronary imaging was used, than with angiography guidance (20.9% vs. 10.4%). This might have occurred because operators in the stepwise provisional group opted to use IVUS or OCT in cases where the procedure was not progressing smoothly.

Even though intravascular imaging use in the EBC MAIN population may be lower than expected given current guideline recommendations(20), it represents real-life current practice, across multiple high volume European sites during the recruitment period (2016 to 2019). In the DKCRUSH-V trial IVUS use was similar at 41%(21). Furthermore, data from 11264 unprotected LM PCIs, from the British Cardiovascular Intervention Society national database, showed that intracoronary imaging was used in 50% of cases in 2014(11). The limited uptake of intracoronary imaging worldwide may be partly due to perceived increased procedural times(22) and costs(23), and inadequate training(24). Indeed, in our study the procedure duration was on average 22 minutes longer among patients who underwent intracoronary imaging, than in those who did not.

There are several trials hoping to clarify the role of imaging in bifurcation coronary disease. The OPTIMAL trial (optimization of left main PCI with intravascular ultrasound, NCT04111770) is randomizing patients to IVUS or angiogram guidance for LM PCI, and is using standardized IVUS criteria that define adequate stent optimization. The DKCRUSH VIII trial (IVUS Guided DK Crush Stenting Technique for Patients with Complex Bifurcation Lesions, NCT03770650) is comparing the composite of cardiac death, MI or TVR at 1-year for an IVUS-guided vs. angiography-guided dual-stent strategy for complex bifurcation lesions, according to DEFINITION criteria(16), including LM bifurcations. The OCTOBER trial (European Trial on Optical Coherence Tomography Optimised Bifurcation Event Reduction; NCT03171311) is comparing 2-year MACE for OCT- vs. angiography-guided bifurcation stenting, in LM or non-LM disease. The ILUMIEN IV trial is comparing OCT- vs. angiography-guided PCI, in complex lesions including non-left-main bifurcations treated with a dual-stent strategy (NCT03507777).

A clear limitation of this study is the fact that patients were not randomly allocated to the use of intravascular imaging. Therefore, there is selection bias inherent in the results we have demonstrated. Additionally, because this study was not principally concerned with intravascular imaging, criteria for optimal stent deployment were not recorded after performing IVUS. It is unclear if adequate stent optimization was achieved in all patients. Future studies are needed to validate and standardize IVUS-guided stent optimization methods. Furthermore, although we had data on whether operators re-intervened based on IVUS findings, we did not have information on whether operators re-intervened based on OCT findings. Importantly, the rates of the primary outcome limited power to detect statistically significant associations. Hence, the study was underpowered to detect significant interactions between intracoronary imaging use, the primary outcome, and additional factors. Finally, although intracoronary imaging use was a pre-specified sub-group, no adjustment for multiplicity was made in this analysis. Due to the potential for type 1 statistical error, the findings of this analysis should be interpreted as exploratory and not definitive.

## Conclusions

In conclusion, in the EBC MAIN trial, the primary outcome did not differ between patients who did or did not have intracoronary imaging, nor for those who had re-intervention based on IVUS findings. However, in patients where no intracoronary imaging was undertaken, clinical outcomes were worse with the systematic dual-stent strategy compared to the provisional approach, and when intracoronary imaging was used, there was a trend toward better clinical outcomes with the systematic dual-stent strategy compared to the provisional approach.

### Impact on daily practice

When angiography is used to guide PCI there can be unrecognized stent under-expansion. Observational studies and subgroup analyses from clinical trials suggest potential benefit from IVUS use during LM PCI, however, the value of IVUS has never been proven in appropriately powered randomized trials. In this study, overall there was no difference in clinical outcomes between intracoronary imaging use and angiogram guidance, during LM bifurcation PCI. However, not using intracoronary imaging may worsen outcomes for dual-stent, compared to single-stent approaches. This suggests that intracoronary imaging might be important to optimize results particularly from dual-stent strategies. Future studies are needed to validate and standardize IVUS-guided stent optimization methods, and future studies comparing IVUS-vs. angiography-guided PCI should include standardized criteria that define adequate stent optimization.

## Data Availability

All data is available in the manuscript

## Abbreviations

IVUS: intravascular ultrasound
LM: Left main
MACE: major adverse cardiac events
MI: myocardial infarction
OCT: optical coherence tomography
OR: odds ratio
PCI: percutaneous coronary intervention
SD: standard deviation
TVR: target vessel revascularization
TIMI: Thrombolysis in Myocardial Infarction

## Acknowledgements

We want to acknowledge and thank the support of the European Bifurcation Club with all its members: Dr Remo Albiero, Dr Adrian Banning, Dr Francesco Burzotta, Dr Yiannis Chatzizisis, Dr Alaide Chieffo, Dr Olivier Darremont, Dr Miroslaw Ferenc, Dr David Hildick-Smith, Dr Tom Jonhson, Dr Jens Lassen, Dr Thierry Lefevre, Dr Yves Louvard, Dr Manual Pan, and Dr Goran Stankovic. We also acknowledge Dr Anno Diegeler, Dr Norbert Frey, Dr Jan Tijssen as DSMB members and Dr Jacques Machecourt, Dr Lisette O. Jensen and Dr Jose R. Rumoroso as CEC members. We thank all the CERC Team, including Dr Marie-Claude Morice, Alice Popineau, Laure Morsiani, Benedicte Borsik, and Dr Antoinette Neylon and her Corelab Team for the analyses performed. We thank all the principle investigators at each site: Dr Adrian Wlodarczak, Dr Adrian Banning, Dr Philippe Brunel, Dr Francesco Burzotta, Dr Alaide Chieffo, Dr Evald Christiansen, Dr Gerald Clesham, Dr David Hildick-Smith, Dr Mohaned Egred, Dr Andrejs Erglis, Dr Evgeny Kretov, Dr Jean Fajadet, Dr Miroslaw Ferenc, Dr Frédéric Bouisset, Dr Thomas Hovasse, Dr René Koning, Dr Krzysztof Reczuch, Dr Marc Silvestri, Dr Monica Masotti, Dr Lindsay Mitchell, Dr Darren Mylotte, Dr Olivier Darremont, Dr Manuel Pan, Dr Pierluigi Omedè, Dr Simon Redwood, Dr Antonieta Serra, Dr Mark Spence, Dr Goran Stankovic, Dr Thomas Enströem, Dr Thomas Schmitz, and Dr Beatriz Vaquerizo. We thank Medtronic for the grant which allowed us to undertaken this trial. Finally, the contribution of all the patients who willingly participated in this study is gratefully acknowledged.

